# Red Blood Cell-Derived miR-93-5p Correlates with PD-1/PD-L1 Upregulation and Poor Prognosis in Lung Cancer

**DOI:** 10.64898/2025.12.11.25342074

**Authors:** Pushpa Dhilipkannah, Feng Jiang

## Abstract

**Background:** Lung cancer remains the deadliest malignancy. Although PD-1/PD-L1 immune checkpoint inhibitors have improved survival, their benefit is limited by persistent immune resistance. Identifying systemic regulators of tumor immune microenvironment may enhance therapeutic efficacy. We previously showed that RBC-derived miR-93-5p promotes PTEN loss and PI3K–AKT activation. This study examined whether RBC-derived miR-93-5p is associated with immune checkpoint activity and features of T-cell exhaustion in lung cancer.

**Methods:** RBCs, RBC-derived exosomes, and plasma were isolated from 80 lung cancer patients and 30 controls, and tumor and matched noncancerous lung tissues were collected. miR-93-5p expression was quantified by droplet digital PCR. PD-1, PD-L1, CD8^+^ T-cell infiltration, CD69, IFN-γ, and TNF-α were evaluated by immunohistochemistry. Associations with clinicopathologic features and survival were statistically analyzed.

**Results:** miR-93-5p levels in RBCs, their exosomes, and tumor tissues were significantly elevated in lung cancer. Higher miR-93-5p was associated with increased PD-1 and PD-L1 expression and reduced CD69, IFN-γ, and TNF-α, consistent with T-cell exhaustion. Elevated miR-93-5p correlated with advanced disease and reduced survival.

**Conclusions:** RBC-derived miR-93-5p is associated with immune checkpoint activation, T-cell exhaustion, and poor clinical outcomes, suggesting impaired antitumor immunity. Targeting this axis may improve the efficacy of immunotherapy in lung cancer.

## Introduction

Non-small cell lung cancer (NSCLC) accounts for approximately 85% of all lung cancer cases and remains the leading cause of cancer-related mortality worldwide ^1^. There are two predominant histological subtypes of NSCLC, adenocarcinoma (AC) and squamous cell carcinoma (SCC), both of which rely significantly on the tumor immune microenvironment (TIME) for their progression ^1^. Immunotherapies targeting programmed death-1 (PD-1) and programmed death-ligand 1 (PD-L1) have transformed NSCLC treatment by restoring antitumor immune activity, resulting in durable clinical responses in a subset of patients ^2^. However, inconsistent clinical responses and immune-related toxicities emphasize the need to elucidate the molecular drivers of immune escape and determinants of PD-1/PD-L1 inhibitor efficacy to ultimately reduce lung cancer mortality ^2^.

We recently uncovered a previously unrecognized role of red blood cells (RBCs) in lung cancer biology^3, 4^. Beyond their traditional role as oxygen carriers, RBCs actively modulate tumor biology by delivering regulatory molecular cargo via RBC-derived exosomes ^3, 4^. Particularly, RBC-derived exosomal miR-93-5p can transfer to tumor cells, where it suppresses PTEN and activates PI3K-AKT signaling, key pathways known to promote oncogenesis and immune evasion^4^. Conversely, tumor-derived exosomes enrich miR-93-5p within RBCs, establishing a bidirectional amplification loop that accelerates tumor progression ^3, 4^. In mouse models, RBC-derived miR-93-5p accelerated tumor growth and reduced survival, whereas antisense inhibition of miR-93-5p or blockade of exosome release/uptake attenuated tumor-promoting phenotypes ^3, 4^. Because PTEN loss and PI3K–AKT activation upregulate PD-L1 and suppress T-cell effector function^5^, we hypothesized that RBC-derived miR-93-5p contributes to an immunosuppressive TIME in NSCLC by enhancing PD-L1 expression and limiting T-cell activity. The objective of this study was to examine whether RBC-derived miR-93-5p is associated with immune checkpoint activity, features of T-cell exhaustion, and clinical outcomes in lung cancer usi ng integrated molecular, immunohistochemical, and survival analyses.

## Materials and Methods

### Patients and clinical specimens

Ethical approval for this study was obtained from the Institutional Review Board of the University of Maryland, Baltimore (IRB approval number: HP-00040666). All procedures involving human participants were conducted in accordance with institutional and federal ethical guidelines. Written informed consent was obtained from all participants prior to enrollment and biospecimen collection. The study was conducted during the IRB-approved period, from April 18, 2022, to April 17, 2023. The cohort included 80 individuals with NSCLC and 30 cancer free smokers (Table 1). Peripheral blood was collected into ethylenediaminetetraacetic acid-coated tubes (BD, Franklin Lakes, NJ) following standard clinical procedures, and processed within two hours ^6-8^. For patients with NSCLC, formalin-fixed, paraffin-embedded (FFPE) tumor tissues and corresponding adjacent non-cancerous lung tissues were obtained through surgical resection or biopsy. Tumor cellularity (≥70%) was confirmed by hematoxylin and eosin (H&E) staining in accordance with WHO and AJCC (8th edition) criteria ^9^. Histological subtypes, including AC and SCC, were classified following the World Health Organization criteria. Staging assessments were further supported by imaging modalities such as computed tomography (CT), positron emission tomography (PET-CT), and magnetic resonance imaging (MRI), together with corresponding pathological reports ^10, 11^. Comprehensive clinical data were curated for all NSCLC patients, including diagnostic details, tumor features, treatment regimens, and coexisting conditions. Longitudinal follow-up was conducted through routine clinical appointments, review of electronic health records, and linkage to the state cancer registry. The follow-up duration ranged from 6 to 60 months. Overall survival (OS) was defined as the time interval from the date of pathological diagnosis to death or last documented follow-up. Vital status and dates of death were validated using hospital databases, death certificates, and the Social Security Death Index.

**Table 1.** Clinical and Demographic Characteristics of 80 NSCLC Patients.

|  |  |
| --- | --- |
| Age (Mean ± SD, years) | 66 (13.7) |
| Sex |  |
| - Female | 25 |
| - Male | 55 |
| Race |  |
| - AA | 23 |
| - WA | 57 |
| Smoking Pack-Years (Median) | 46 |
| Stage |  |
| - Stage I | 43 |
| - Stage II | 30 |
| - Stage III-IV | 7 |
| Histological Type |  |
| - AC | 45 |
| - SCC | 35 |
| NSCLC, non-small cell lung cancer; AA, African Americans; WA, White Americans; AC, adenocarcinoma; SCC, squamous cell carcinoma; SD, standard deviation. |  |

### Preparation of Plasma, RBCs, and RBC-Derived Exosomes

Plasma, RBCs, and RBC-derived exosomes were isolated following the Minimal Information for Studies of Extracellular Vesicles (MISEV) guidelines and our established protocols ^3, 4, 6-8, 12-14^. Plasma was obtained by centrifugation at 1,500 × g for 10 min and clarified at 2,000 × g for 15 min at 4 °C, and hemolyzed samples—identified by A414 hemoglobin levels, the miR-451/23a ratio, and plasma haptoglobin—were excluded. RBC-derived exosomes were isolated from plasma using qEVoriginal 70 nm size-exclusion chromatography (Izon Science); sequential fractions were collected, and exosome-enriched fractions were defined by Nanoparticle Tracking Analysis (NTA)-confirmed particle size and concentration with low A280 protein absorbance, then pooled for downstream analyses. After plasma removal, the buffy coat was discarded to obtain purified RBCs, which were washed twice in PBS, lysed in hypotonic buffer (10 mM Tris-HCl, pH 7.4), and centrifuged at 300 × g and 16,500 × g to remove debris. The resulting supernatant was ultracentrifuged at 100,000 × g for 70 min (Optima XE-90; Beckman Coulter), and the exosome pellet was washed, resuspended in PBS, and characterized by NTA and electron microscopy to verify vesicle size distribution and purity.

### Droplet Digital PCR (ddPCR)

Expression levels of miR-93-5p in RBCs, RBC-derived exosomes, plasma, and tissues were quantified using ddPCR with specific primers and probes as described in our previous reports ^3, 4, 15^. Briefly, RNA was first extracted from the specimens using a column-based method (miRNeasy Mini Kit, Qiagen, Germantown, MD) according to the manufacturer’s protocol. RNA was then reverse transcribed using the TaqMan MicroRNA Reverse Transcription Kit (Thermo Fisher Scientific, Waltham, MA) for miRNAs. Each 20 µL reaction contained 2× ddPCR Supermix for Probes (no dUTP) (Bio-Rad, Hercules, CA), 250 nM TaqMan probe, 900 nM primers, and 2 µL cDNA. Droplets were generated on a QX100 Droplet Generator and amplified (95 °C 10 min; 40 cycles of 94 °C 30 s and 60 °C 1 min; 98 °C 10 min). Droplet fluorescence was analyzed using a QX100 Droplet Reader and QuantaSoft Software (Bio-Rad). miR-451a was used as the endogenous reference miRNA for the RBC and exosomal assays. Absolute copy numbers were calculated using Poisson statistics.

### Immunohistochemical (IHC)

Immunohistochemistry (IHC) was performed on FFPE tissue sections to evaluate PD-1, PD-L1, CD8^+^ T-cell infiltration, CD69, IFN-γ, and TNF-α expression using validated primary antibodies as previously described. ^4, 16, 17^ (Supplementary Table 1). Briefly, sections were deparaffinized, rehydrated, and subjected to heat-induced antigen retrieval in citrate buffer prior to antibody incubation. Primary antibodies (Abcam, Waltham, MA) were applied according to the manufacturers’ instructions. Sections were then incubated with species-specific horseradish peroxidase (HRP)-conjugated secondary antibodies (goat anti-mouse IgG-HRP or goat anti-rabbit IgG-HRP; Jackson ImmunoResearch, West Grove, PA). Signal detection was achieved through chromogenic development, and slides were counterstained with hematoxylin. Images were captured using an Olympus light microscope (Center Valley, PA). For quantitative evaluation, five representative fields containing both tumor and stromal regions were randomly selected from each specimen. Positively stained cells exhibiting distinct brown chromogenic labeling were counted manually using AnalySIS Pro software (Olympus). The percentage of positive cells per 100 counted was converted into an ordinal scoring system: 0 (0-10%), 1 (11-25%), 2 (26-50%), and 3 (>50%) ^16, 18, 19^. Each region was independently assessed by two investigators in triplicate under blinded conditions to ensure consistency and eliminate observer bias.

### Statistical Analysis

To achieve a statistical power of 85% for detecting clinically meaningful group differences (α = 0.05), we enrolled 80 patients with lung cancer and 30 cancer-free controls. All statistical procedures were conducted using SAS 9.4 (SAS Institute, Cary, NC). Data normality was assessed by the Shapiro-Wilk test and Q-Q plots. Group comparisons used Student’s t-test or Mann-Whitney U test for continuous variables and Chi-square or Fisher’s exact test for categorical data. Correlations between miR-93-5p and immune markers were examined using Pearson or Spearman coefficients. Overall survival was evaluated by Kaplan-Meier and log-rank tests, and independent predictors were identified by Cox regression. Variables with p < 0.10 or biological relevance entered multivariate models. Statistical significance was defined as p < 0.05. Graphs were generated in GraphPad Prism 10.0.

## Results

### Altered RBC-miR-93-5p Levels and Immune Checkpoint Activity in Lung Cancer

Consistent upregulation of miR-93-5p was observed in RBCs and RBC-derived exosomes from lung cancer patients relative to cancer-free controls (all p < 0.05; Table 2, Figure 1). In contrast, plasma miR-93-5p levels showed no significant difference between the two groups (Figure 1). Furthermore, the expression level of miR-93-5p was significantly higher in lung tumor tissues compared with the matched noncancerous lung tissues (p < 0.05; Table 2, Figure 1). In addition, a correlation was observed for miR-93-5p expression levels across RBCs, RBC-derived exosomes, and lung tumor tissues in lung cancer patients (r = 0.87-0.91, p < 0.05; Figure 2).

**Table 2.** Expression levels of miR-93-5p and immune markers in lung cancer patients and controls.

|  |  |  |  |
| --- | --- | --- | --- |
| **Markers** | **Cancer Patients OR Lung Tumor Tissues (Mean ± SD)** | **Controls or Matched Noncancerous Lung Tissues (Mean ± SD)** | **p-value** |
| miR-93-5p in RBCs * | 50.68 ± 8.76 | 35.39 ± 7.93 | 0.017 |
| miR-93-5p in RBC-derived exosomes * | 53.48 ± 8.76 | 39.03 ± 10.02 | 0.026 |
| miR-93-5p in plasma * | 11.26 ± 3.52 | 11.48 ± 3.09 | 0.894 |
| miR-93-5p in lung tissues # | 61.89 ± 10.38 | 41.87 ± 10.53 | <0.001 |
| PD-L1 (% positive cells) # | 27.481 ± 7.487 | 24.769 ± 4.823 | 0.045 |
| PD-1 (% positive cells) # | 21.498 ± 6.476 | 18.032 ± 4.013 | 0.022 |
| CD8<sup>+</sup> T cells (% positive cells) # | 16.024 ± 4.012 | 15.889 ± 4.682 | 0.658 |
| CD69 (% positive cells) # | 8.013 ± 2.095 | 14.478 ± 2.599 | 0.037 |
| IFN-γ (% positive cells) # | 10.012 ± 3.981 | 19.041 ± 3.789 | 0.046 |
| TNF-α (% positive cells) # | 15.017 ± 3.002 | 27.021 ± 5.986 | 0.039 |

\* miR-93-5p levels in RBCs and RBC-derived exosomes were compared between 80 lung cancer patients and 30 cancer-free controls. # Tissue miR-93-5p and all immune marker analyses were based on paired tumor and matched noncancerous lung tissues from the same 80 patients.

**Figure 1.**
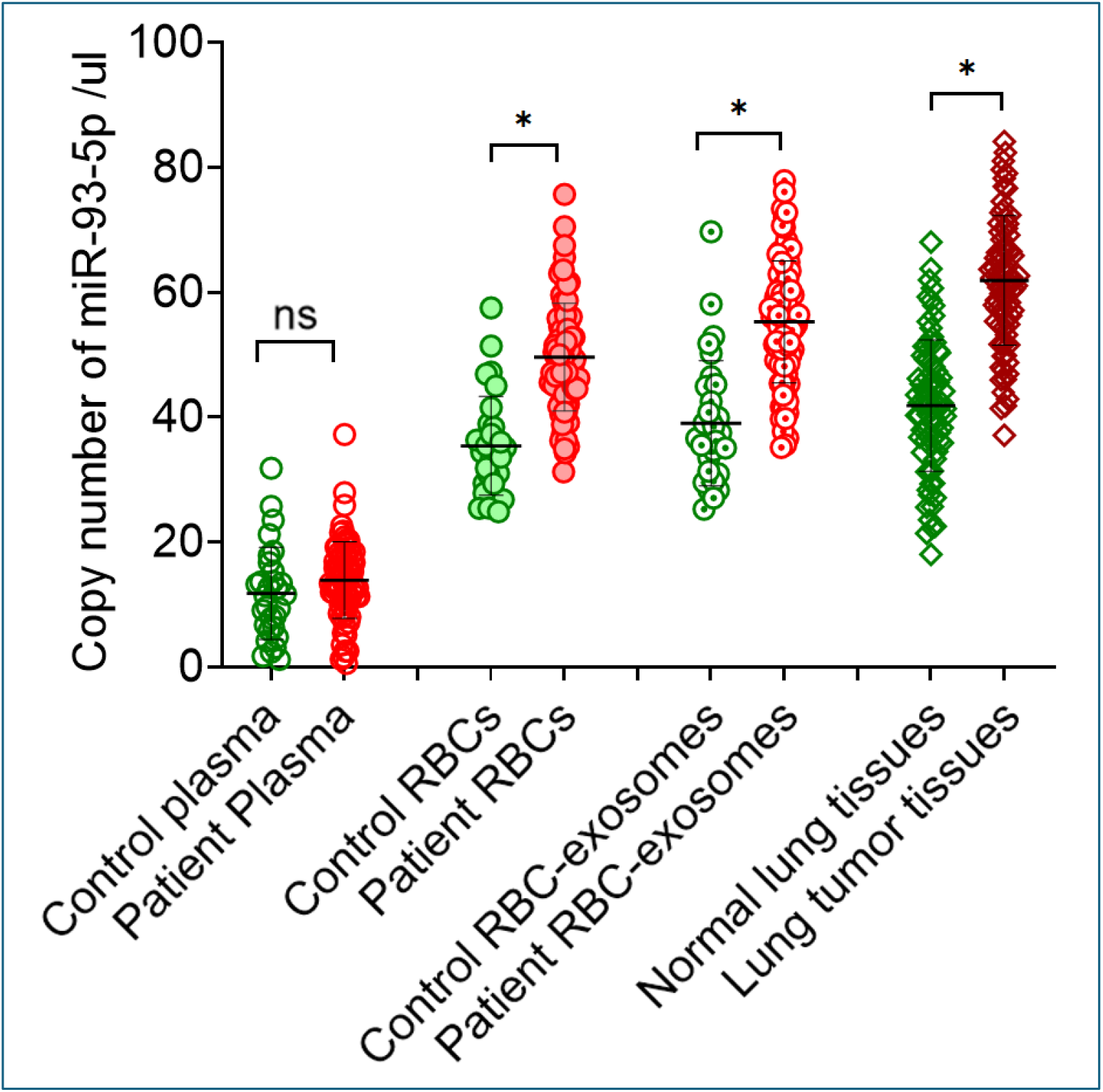
Expression analysis of miR-93-5p in plasma, RBCs, RBC-derived exosomes, and lung tumor tissues. miR-93-5p levels were significantly elevated in RBCs and RBC-derived exosomes from lung cancer patients relative to cancer-free controls (p=0.002), whereas plasma levels showed no significant difference between groups (p=0.001). In lung tumor tissues of 80 patients, miR-93-5p expression was significantly higher than in the matched noncancerous lung tissues (p=0.001). Statistical comparisons were performed using the Mann-Whitney U test for unpaired samples and the Wilcoxon signed-rank test for paired tumor and noncancerous tissues. Asterisks (*) indicate significant differences, and ns denotes non-significant differences.

**Figure 2.**
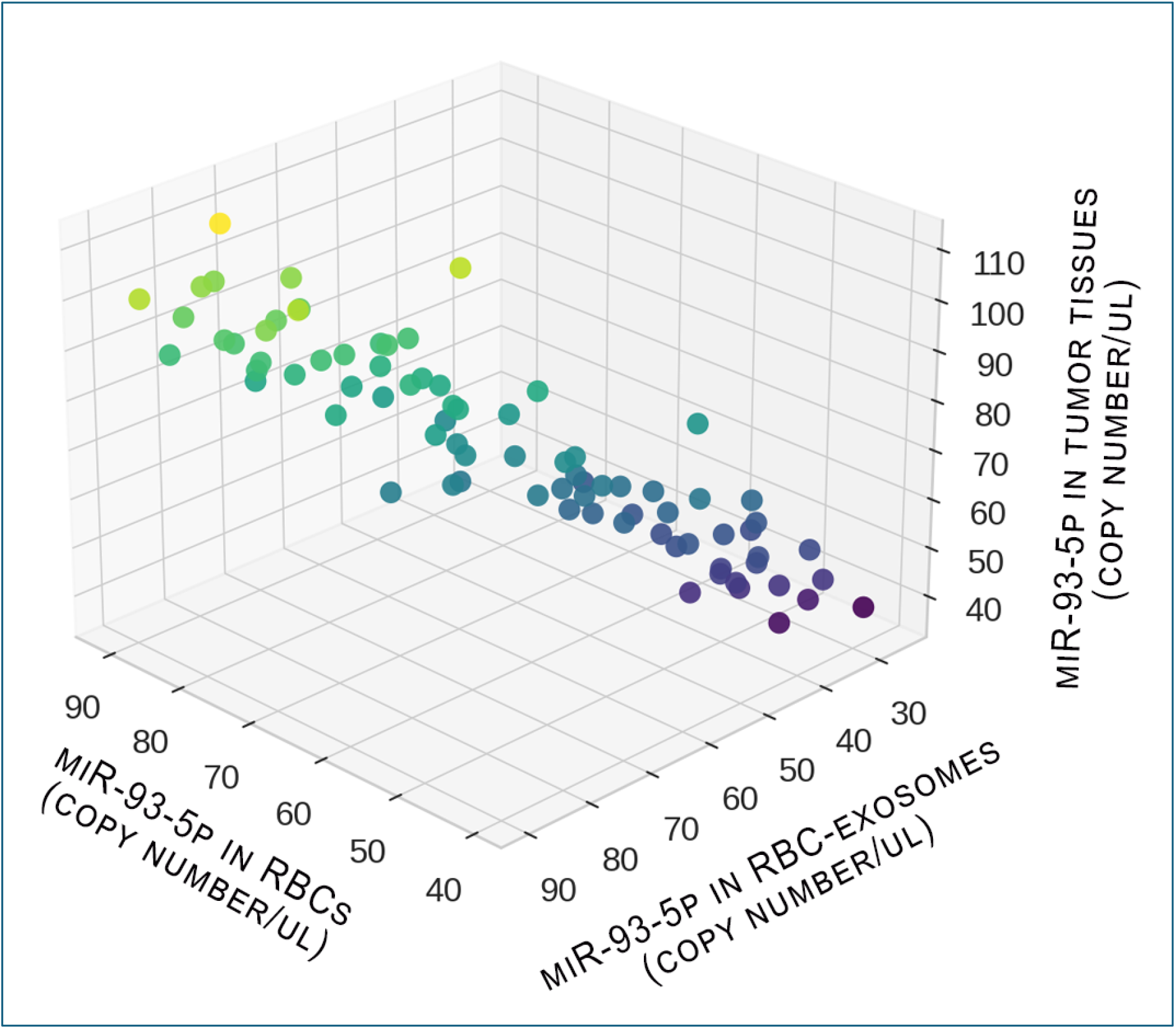
Three-dimensional scatterplots showing correlations of miR-93-5p expression among RBCs, RBC-derived exosomes, and lung tumor tissues from 80 matched cases. Each dot represents an individual patient sample and is color-coded according to the absolute miR-93-5p copy number per µL. Pearson correlation analysis revealed strong positive associations between RBCs and exosomes (r = 0.87). RBCs and tumor tissues (r = 0.91), and exosomes and tumor tissues (r = 0.87). All p<0.05.

Immunohistochemical analyses of PD-1, PD-L1, CD8^+^ T-cell infiltration, CD69, IFN-γ, and TNF-α were successfully performed on the tissue sections (Figure 3). The expression levels of PD-L1 and PD-1 were significantly elevated in tumor tissues, whereas CD69, IFN-γ, and TNF-α levels were markedly reduced (all p < 0.05) (Table 2). However, the proportion of CD8^+^ T cells did not differ significantly between tumor and adjacent normal tissues (p > 0.05) (Table 2).

**Figure 3.**
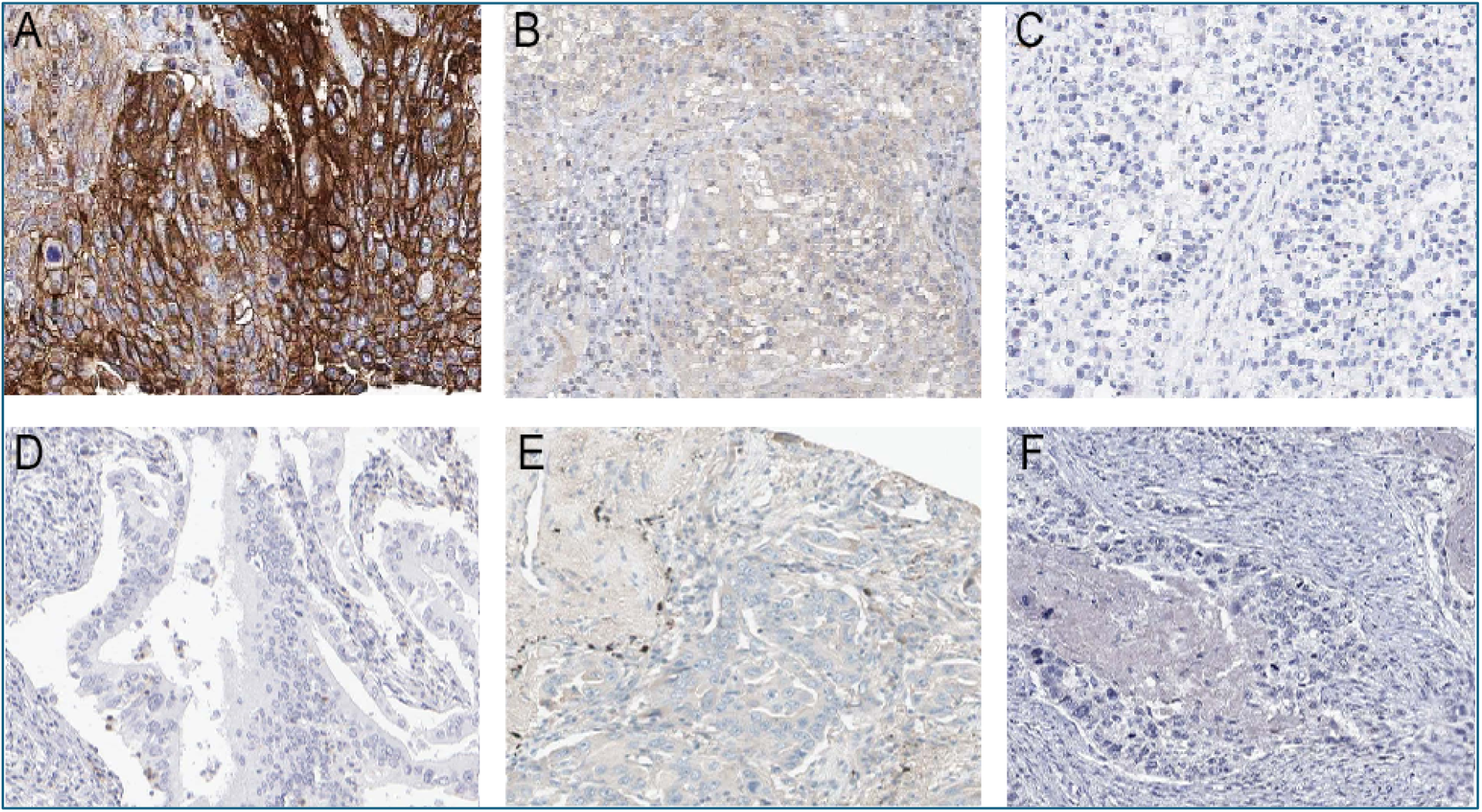
Representative immunohistochemical staining images for immune markers in lung cancer tissues. (A) Strong membranous PD-L1-positive staining in tumor cells of squamous cell carcinoma (SCC). (B) Weak to moderate membranous PD-1 positivity on tumor-infiltrating lymphocytes in adenocarcinoma (AC). (C) CD8^+^ T cells showing distinct cytoplasmic positivity, indicating infiltration in AC. (D) CD69-positive lymphocytes with weak cytoplasmic staining; tumor cells are negative in AC. (E) IFN-γ staining is largely negative, with only rare faint cytoplasmic positivity in infiltrating immune cells in AC. (F) Weak cytoplasmic TNF-α positivity in tumor-infiltrating immune cells, whereas tumor cells in SCC remain negative. All images were acquired at 200× magnification.

### miR-93-5p Correlates with Immune Signatures in Lung Cancer

Pearson’s correlation analysis showed that elevated miR-93-5p levels in RBCs, RBC-derived exosomes, and tumor tissues were significantly positively correlated with increased expression of the immune checkpoint molecules PD-L1 and PD-1 (all p < 0.05; Supplementary Table 2; Figure 4). In contrast, miR-93-5p levels demonstrated significant inverse correlations with the T-cell activation marker CD69 and with the effector cytokines IFN-γ and TNF-α across all specimen types (all p < 0.05). Correlations with CD8^+^ T-cell infiltration were negative but did not reach statistical significance (p > 0.05). Together, these findings indicate that upregulation of miR-93-5p in RBCs, their exosomes, and tumor tissues is associated with enhanced immune checkpoint signaling and reduced antitumor T-cell activity, reflecting an immunosuppressive phenotype in NSCLC.

**Figure 4.**
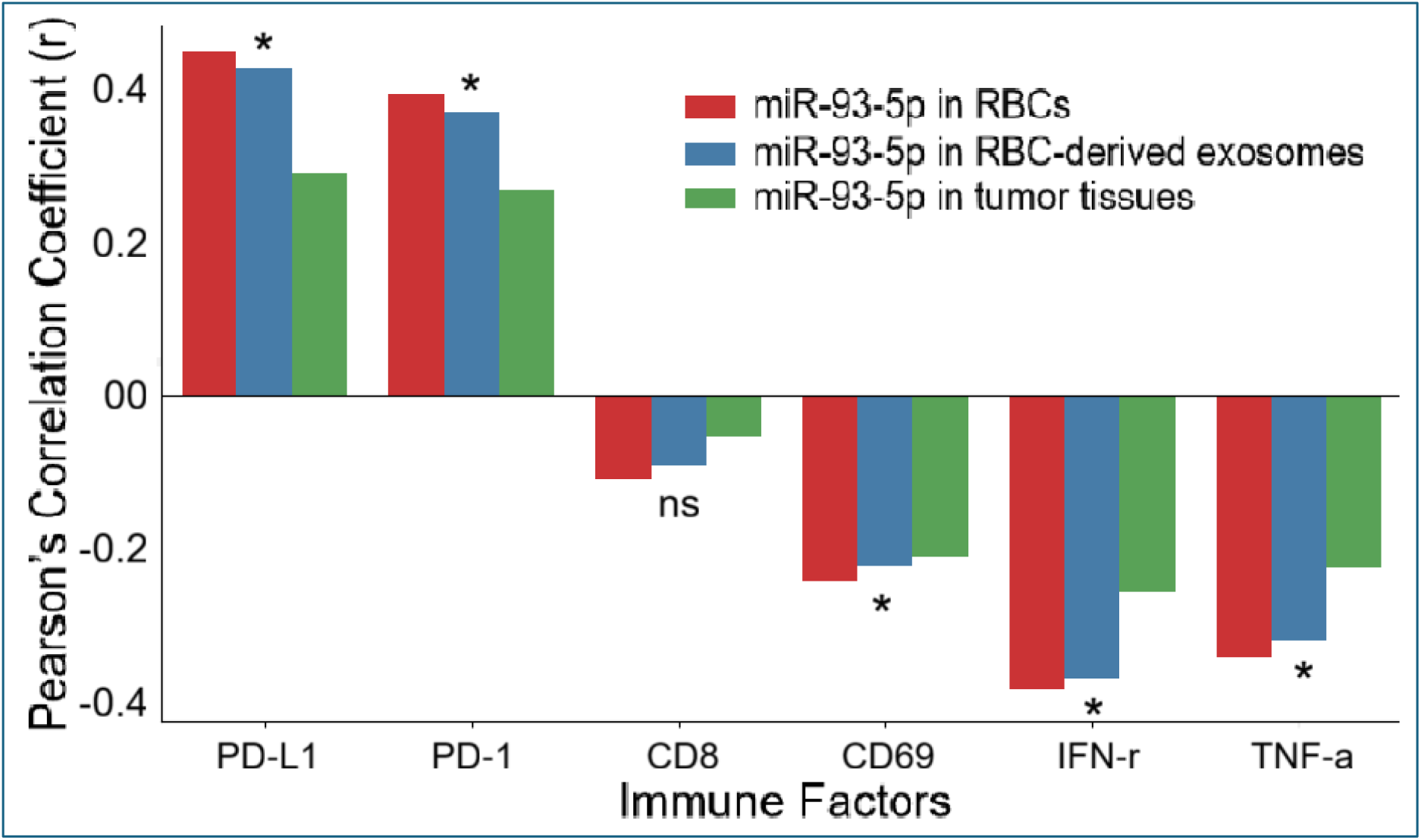
Correlations between miR-93-5p expression and immune regulatory markers in NSCLC. Pearson correlation coefficients (r) are shown for miR-93-5p levels in RBCs (red), RBC-derived exosomes (blue), and tumor tissues (green) in relation to PD-L1, PD-1, CD8^+^ T cells, CD69, IFN-γ, and TNF-α. miR-93-5p exhibited positive correlations with PD-L1 and PD-1 and negative correlations with CD69, IFN-γ, and TNF-α across compartments, whereas correlations with CD8^+^ T cells were weak and not significant. Values correspond to results summarized in Table 3. Asterisks (*) indicate significant differences, and ns denotes non-significant differences.

### Clinical Significance of miR-93-5p and Immune Factors in Lung Cancer Patients

To evaluate the prognostic value of molecular alterations, we performed a multivariate Cox proportional hazards regression analysis (Supplementary Table 3; Figure 5). Higher miR-93-5p expressions in RBCs, RBC-derived exosomes, and tumor tissues were independently associated with an increased risk of mortality, with hazard ratios ranging from 1.78 to 2.10 (all p<0.05). Among immune markers, elevated PD-L1 and PD-1 expressions were significant independent predictors of worse prognosis (HR = 1.847 and 1.648, respectively; all p<0.05). In contrast, CD69, IFN-γ, and TNF-α did not reach statistical significance in the multivariate model. Among clinical factors, advanced disease stage exhibited the association with poor survival (HR = 3.002; p = 0.0373). Age and gender were not associated with survival (Supplementary Table 3; Figure 5).

**Figure 5.**
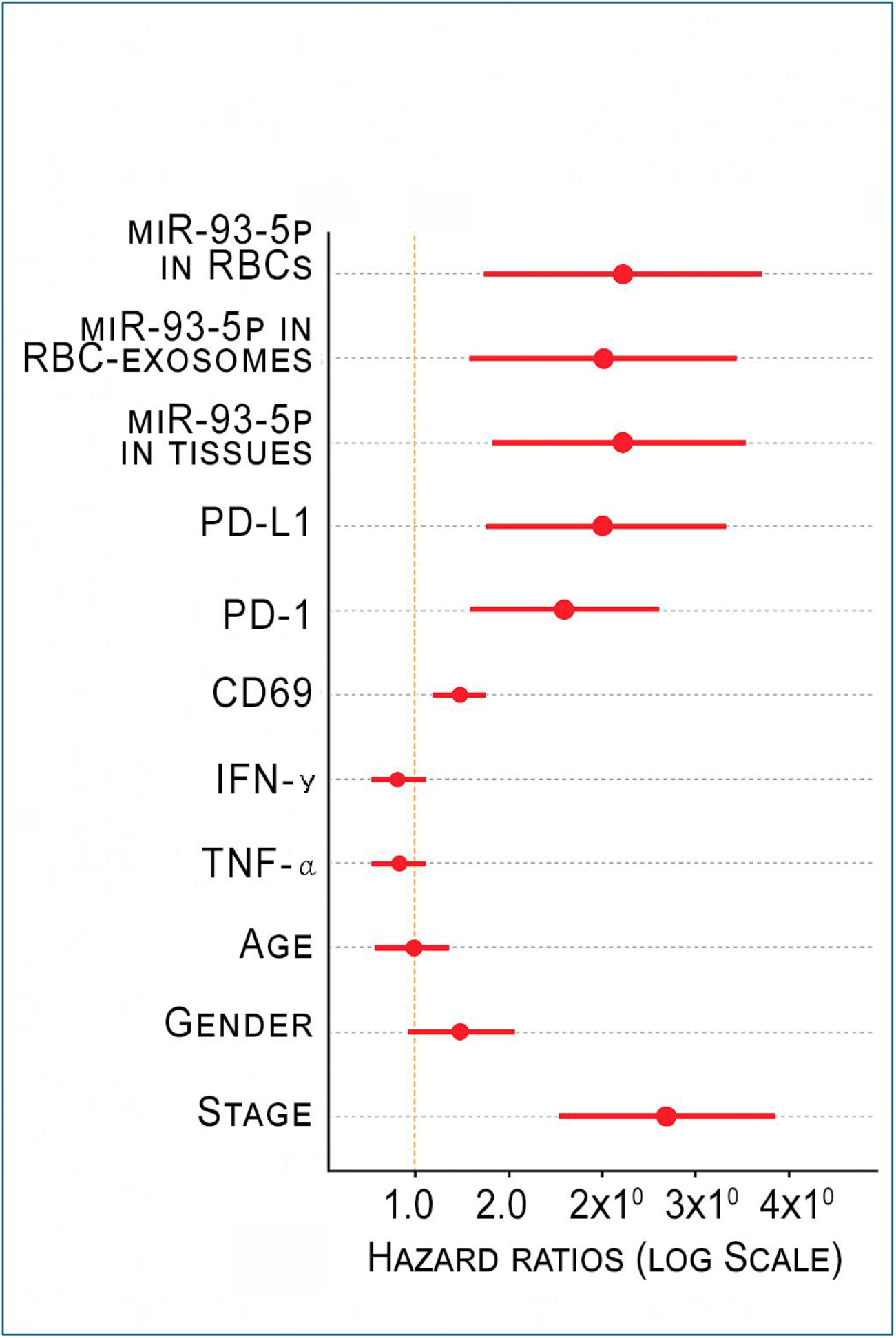
Forest plot summarizing hazard ratios (HRs) and 95% confidence intervals (CIs) derived from a multivariate Cox proportional hazards regression model assessing the impact of miR-93-5p expression, immune markers, and clinical variables on patient outcomes. HRs are shown on a log scale with a vertical reference line at HR = 1. Elevated miR-93-5p levels in RBCs, RBC-derived exosomes, and tumor tissues, along with higher PD-L1 and PD-1 expressions, were significantly associated with increased hazard. CD69, IFN-γ, TNF-α, and age demonstrated borderline associations, whereas gender showed no significant effect. Advanced disease stage (III-IV vs. I-II) exhibited the strongest association with poorer survival.

To further characterize the biological and clinical relevance of the molecules, we performed univariate association analyses to examine the relationships between miR-93-5p expression and immune factors, clinical characteristics, and overall survival. As shown in Supplementary Table 4, elevated miR-93-5p levels in RBCs, RBC-derived exosomes, and tumor tissues were significantly correlated with advanced disease stage and poor overall survival (p < 0.05). Patients with high miR-93-5p expression exhibited markedly shorter survival times than those with low expression, supporting the link between miR-93-5p upregulation, aggressive tumor behavior, and unfavorable prognosis. miR-93-5p expression did not differ by age, sex, or histological subtype (AC vs. SCC), indicating that its prognostic impact is independent of these factors.

Moreover, we evaluated whether integrating miR-93-5p expression across multiple compartments, RBCs, RBC-derived exosomes, and tumor tissues, could improve prognostic performance beyond individual measurements. Consistent with our earlier finding that each compartment-specific measure of miR-93-5p was independently prognostic. The integrated model failed to enhance overall survival discrimination compared with single-compartment analyses, as demonstrated by multivariate Cox regression and corresponding survival curves (Figure 6; Supplementary Table 5).

**Figure 6.**
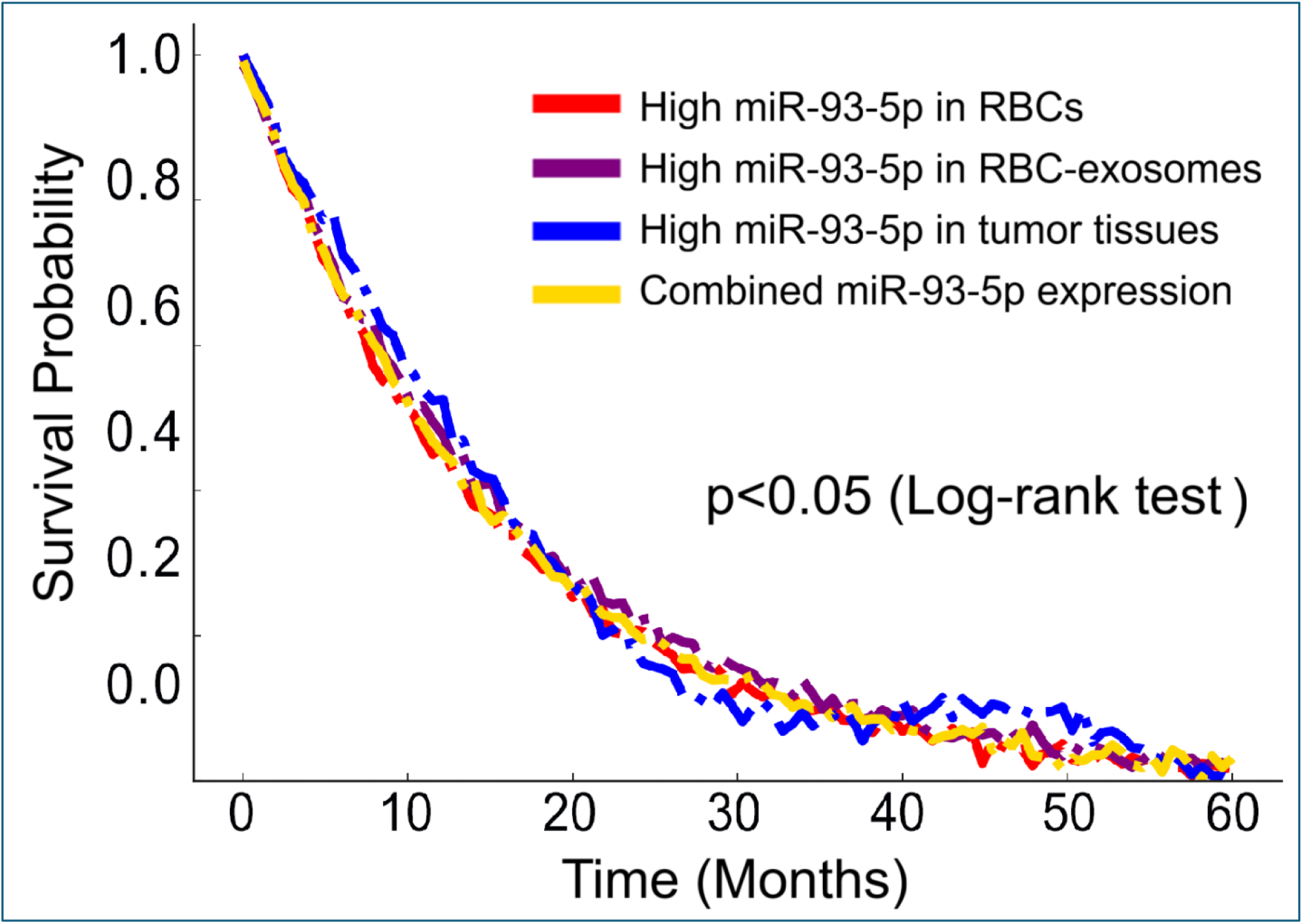
Kaplan-Meier survival curves illustrating the prognostic relevance of miR-93-5p expression in lung cancer patients. Each line represents high miR-93-5p levels in a specific compartment: RBCs (red), RBC-derived exosomes (purple), and lung tumor tissues (blue), as well as combined expression across all compartments (yellow). The curves display mild divergence and natural crossing, indicating biological variability but no statistically significant survival differences among groups (p > 0.05, log-rank test). Overall, similar slopes and endpoints suggest that elevated miR-93-5p expression across compartments correlates with comparable clinical outcomes, highlighting its consistent prognostic pattern in lung cancer.

## Discussion

This study demonstrates that miR-93-5p is upregulated in RBCs, RBC-derived exosomes, and lung tumor tissues, and this elevation is closely associated with dysregulation of immune checkpoint pathways in NSCLC. Particularly, the increased miR-93-5p levels consistently correlated with elevated PD-1/PD-L1 expression and reduced CD69, IFN-γ, and TNF-α levels, indicative of impaired T-cell activation. These molecular alterations were further linked to advanced disease stage and reduced survival. Therefore, RBC-associated miR-93-5p may serve as a mechanistic bridge between systemic molecular signaling and local immune suppression within the TIME, thereby contributing to NSCLC development and progression.

As the most lethal cancer worldwide, NSCLC demands more effective treatment options^1^. Although PD-1/PD-L1 inhibitors have transformed cancer therapy, major challenges persist in lung cancer due to wide interpatient variability in response and immune-related toxicities ^2^. Prior studies indicate that TIME composition strongly influences responsiveness to immune checkpoint blockade, yet the upstream molecular regulators that shape these immune contexts are not fully defined ^20-25^. Our findings provide new insight by demonstrating that, in addition to tumor-derived miRNAs, a circulating miRNA reservoir originating from RBCs correlates with key immune checkpoint pathways in NSCLC. This systemic source may help explain why some tumors demonstrate heightened PD-L1 expression and T-cell dysfunction even before therapy initiation. Importantly, the strong correlation between RBC-derived miR-93-5p and PD-1/PD-L1 upregulation suggests biological and clinical relevance. We previously demonstrated that RBC-derived exosomal miR-93-5p is transferred into lung cancer cells, where it suppresses PTEN and activates PI3K–AKT signaling, thereby promoting tumor progression ^4^. Because PTEN loss is known to upregulate PD-L1 through PI3K–AKT signaling ^5^, the associations observed in the present study suggest that elevated RBC-miR-93-5p may contribute to immune escape by enhancing PD-L1 expression and promoting PD-1–mediated T-cell inhibition. Moreover, the associations between elevated miR-93-5p levels, advanced stage, and poor survival underscore its potential as a non-invasive biomarker for disease aggressiveness and immunotherapy stratification. Particularly, RBCs, unlike plasma, offer a stable and long-lived compartment for miRNA accumulation, making them especially attractive for biomarker applications.

Our findings also complement and extend previous observations implicating microRNAs in tumor immune evasion^26-29^. For example, miR-21, miR-155, and miR-34a are known regulators of PI3K/AKT, JAK/STAT, and PD-L1 expression in NSCLC ^26, 27^. miR-93-5p has also been associated with PD-L1 upregulation in breast and colorectal cancer ^28, 29^, but its role in lung cancer immune biology has remained unclear. Here, we reveal that miR-93-5p is not only elevated within the tumor but also enriched systemically in RBCs, which serve as a systemic source influencing oncogenic and immunoregulatory pathways. This extra-tumoral origin underscores a broader and previously underrecognized mechanism through which dysregulated circulating miRNAs influence the TIME and contribute to immune evasion.

We previously demonstrated that antisense inhibition of miR-93-5p or pharmacologic disruption of exosome trafficking resulted in significant tumor regression and improved survival in preclinical models ^4^. The findings of the present study further suggest that targeting RBC-derived miR-93-5p could complement PD-1/PD-L1 blockade by mitigating immune checkpoint activation and T-cell dysfunction, thereby potentially enhancing therapeutic efficacy. For instance, dual targeting, miR-93-5p inhibition plus PD-1/PD-L1 blockade, may represent a rational combination strategy to overcome immune resistance and restore T-cell effector function.

Despite the strengths of this study, additional mechanistic insights are needed. For instance, functional assays assessing T-cell cytotoxicity, cytokine production, or exhaustion states following modulation of RBC-derived miR-93-5p would clarify causal relationships. Spatial multi-omics or single-cell profiling of tumor and immune compartments could further map how RBC-derived signals shape cellular interactions within the TIME. Expanding the cohort size and validating findings across independent patient populations will facilitate clinical translation. Finally, testing miR-93-5p-targeted therapeutics in immunocompetent preclinical models will be crucial for establishing the feasibility of combination approaches with PD-1/PD-L1 inhibitors.

## Conclusion

Elevated RBC-derived miR-93-5p is associated with increased PD-1/PD-L1 activity, features of T-cell exhaustion, and poor prognosis in lung cancer, indicating impaired antitumor immunity. These findings suggest that RBC-derived miR-93-5p functions as a systemic regulator of the TIME and a potential non-invasive biomarker of immune suppression and immunotherapy responsiveness. Furthermore, targeting the RBC-miR-93-5p axis may therefore represent a novel strategy to enhance the efficacy of PD-1/PD-L1 blockade in lung cancer.

## Data Availability

All data produced in the present work are contained in the manuscript

## Supplementary File

**Supplementary Table 1.**
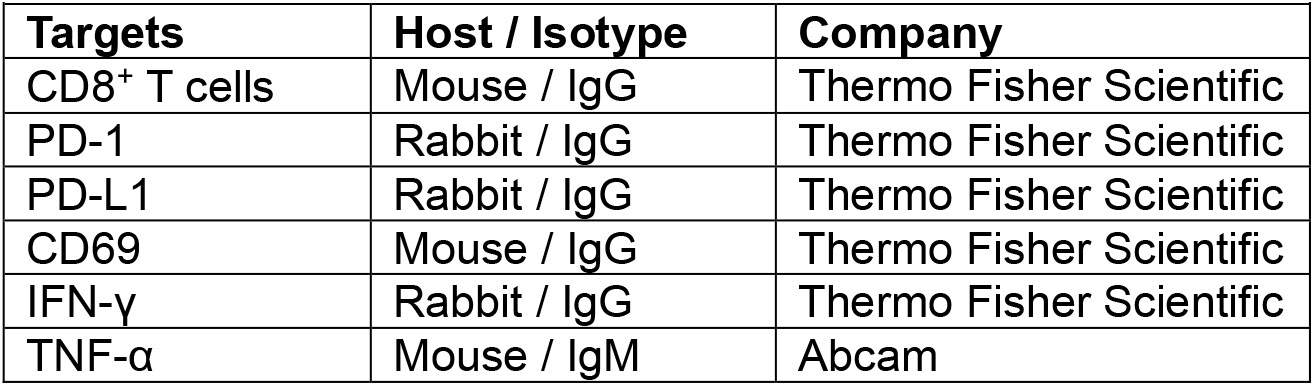
Primary Antibodies Used for IHC.

**Supplementary Table 2.** Pearson’s Correlation Coefficients between miR-93-5p and Immune Factors in Lung Cancer Patients.

|  |  |  |  |  |  |  |
| --- | --- | --- | --- | --- | --- | --- |
| **miR-93-5p** | **PD-L1** | **PD-1** | **CD8** | **CD69** | **IFN-γ** | **TNF-α** |
| In RBCs | $r = +0.482^*$ | $r = +0.421^*$ | $r = -0.294$ | $r = -0.261^*$ | $r = -0.412^*$ | $r = -0.367^*$ |
| In RBC-derived exosomes | $r = +0.458^*$ | $r = +0.396^*$ | $r = -0.275$ | $r = -0.239^*$ | $r = -0.398^*$ | $r = -0.346^*$ |
| In tumor tissues | $r = +0.312^*$ | $r = +0.287^*$ | $r = -0.203$ | $r = -0.228^*$ | $r = -0.276^*$ | $r = -0.241^*$ |

\*Statistically significant correlations ( $p < 0.05$ ).

**Supplementary Table 3.** Multivariate Cox Proportional Hazards Regression Analysis of miR-93-5p Expression and Immune Factors, and Stags.

| **Supplementary Table 3. Multivariate Cox Proportional Hazards Regression Analysis of miR-93-5p Expression and Immune Factors, and Stages** |  |  |  |
| --- | --- | --- | --- |
| **Variable** | **Hazard Ratio** | **95% Confidence Interval** | **p-value** |
| miR-93-5p in RBCs | 1.923 | 1.218 - 3.038 | 0.016 |
| miR-93-5p in RBC-derived exosomes | 1.784 | 1.153 - 2.911 | 0.021 |
| miR-93-5p in tumor tissues | 2.103 | 1.327 - 3.498 | 0.011 |
| PD-L1 | 1.847 | 1.152 - 3.003 | 0.018 |
| PD-1 | 1.648 | 1.104 - 2.503 | 0.025 |
| CD69 | 1.123 | 0.972 - 1.298 | 0.063 |
| IFN-γ | 0.943 | 0.893 - 0.994 | 0.082 |
| TNF-α | 0.911 | 0.872 - 0.963 | 0.073 |
| Age (years) | 1.023 | 1.002 - 1.041 | 0.083 |
| Gender (Male vs. Female) | 1.104 | 0.748 - 1.602 | 0.653 |
| Disease Stage (III-IV vs. I-II) | 3.002 | 1.802 - 5.003 | 0.037 |

Hazard ratios were estimated using multivariate Cox regression.

**Supplementary Table 4.**
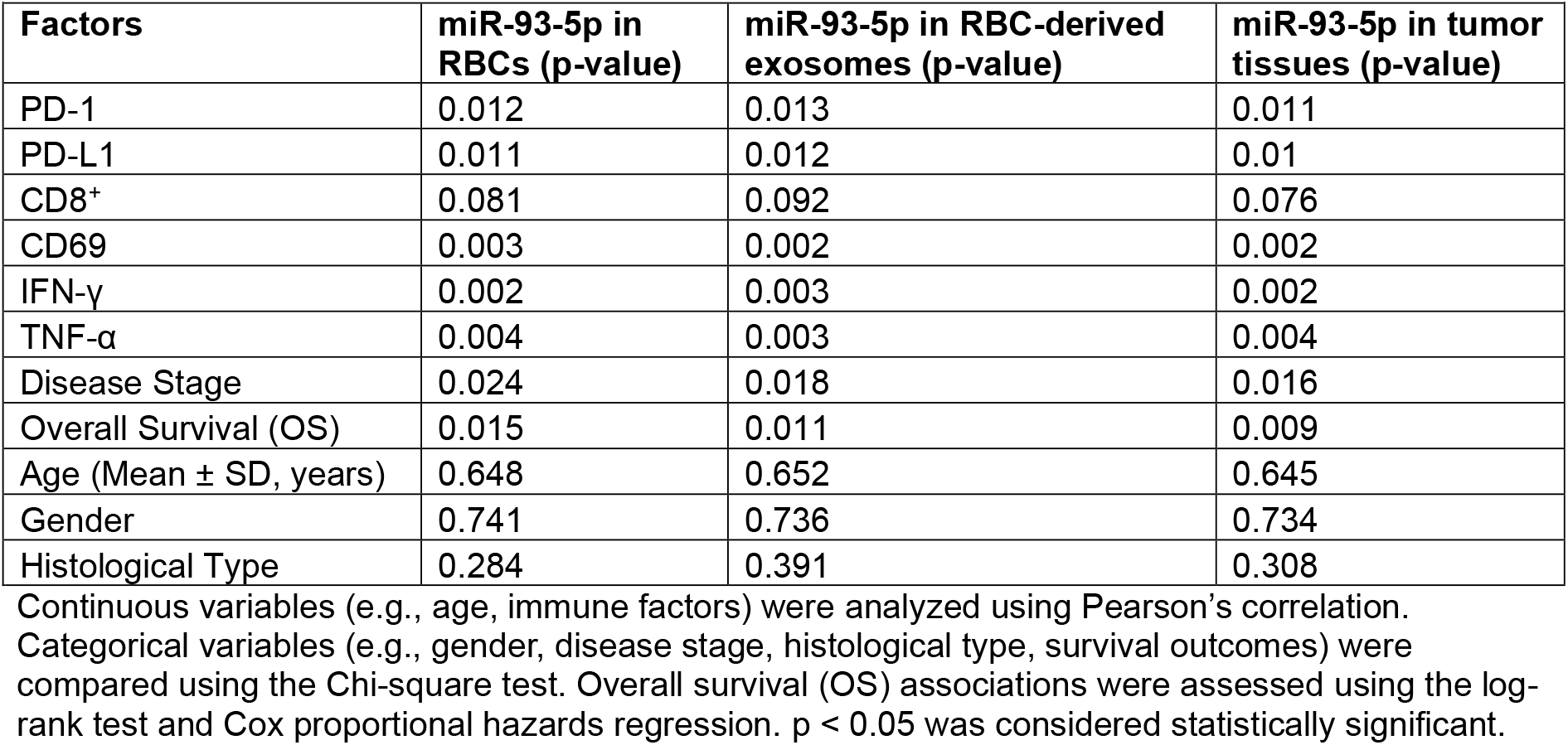
Associations of miR-93-5p Expression Levels with Immune Factors, Clinical Characteristics, and OS.

**Supplementary table 5.** Multivariate Cox Proportional Hazards Regression Analysis of Combined miR-93-5p expression across multiple compartments on Overall Survival (OS)

|  |  |  |  |
| --- | --- | --- | --- |
| **Variables** | **Hazard Ratio (HR)** | **95% Confidence Interval (CI)** | **p-value** |
| miR-93-5p in RBCs | 1.923 | 1.218 - 3.038 | 0.016 |
| miR-93-5p in RBC-derived exosomes | 1.784 | 1.153 - 2.911 | 0.021 |
| miR-93-5p in tumor tissues | 2.103 | 1.327 - 3.498 | 0.011 |
| Combined miR-93-5p expressions | 2.873 | 1.427 - 4.639 | 0.015 |

## Ethics Approval and Consent to Participate

The study was conducted under protocols approved by the Institutional Review Board of the University of Maryland Baltimore (IRB HP-00040666), and all patients provided written informed consent.

## Data Availability

The datasets generated during this study are not publicly available due to patient privacy protections but are available from the corresponding author upon reasonable request.

## Author Contributions

Pushpa Dhilipkannah and Feng Jiang performed experiments and wrote the main manuscript. Pushpa Dhilipkannah1collected samples for this study. Feng Jiang developed the main project idea and conceptual framework. All authors reviewed the manuscript.

## Acknowledgements

The authors wish to thank the Biostatistics Shared Service at the University of Maryland Marlene and Stewart Greenebaum Cancer Center for their invaluable contribution in conducting the statistical analysis for this study.

## References

1 Jonas DE, Reuland DS, Reddy SM et al. Screening for Lung Cancer With Low-Dose Computed Tomography: Updated Evidence Report and Systematic Review for the US Preventive Services Task Force. JAMA 2021; 325 (10): 971–987.

2 Dermani FK, Samadi P, Rahmani G et al. PD-1/PD-L1 immune checkpoint: Potential target for cancer therapy. J Cell Physiol 2019; 234 (2): 1313–1325.

3 Geng X, Ma J, Dhilipkannah P et al. MicroRNA Profiling of Red Blood Cells for Lung Cancer Diagnosis. Cancers (Basel) 2023; 15 (22).

4 Li N, Dhilipkannah P, Holden VK et al. Red Blood Cell-Derived Exosomal miR-93-5p Promotes Lung Cancer Progression through PTEN Suppression. Adv Sci (Weinh) 2025: e11940.

5 Cetintas VB, Batada NN. Is there a causal link between PTEN deficient tumors and immunosuppressive tumor microenvironment? J Transl Med 2020; 18 (1): 45.

6 Li P, Kaslan M, Lee SH et al. Progress in Exosome Isolation Techniques. Theranostics 2017; 7 (3): 789–804.

7 Baranyai T, Herczeg K, Onodi Z et al. Isolation of Exosomes from Blood Plasma: Qualitative and Quantitative Comparison of Ultracentrifugation and Size Exclusion Chromatography Methods. PLoS One 2015; 10 (12): e0145686.

8 Diehl JN, Ray A, Collins LB et al. A standardized method for plasma extracellular vesicle isolation and size distribution analysis. PLoS One 2023; 18 (4): e0284875.

9 Lancia A, Merizzoli E, Filippi AR. The 8(th) UICC/AJCC TNM edition for non-small cell lung cancer staging: getting off to a flying start? Ann Transl Med 2019; 7 (Suppl 6): S205.

10 Del Aguila Mejia J, Armon S, Campbell F et al. Understanding the use of evidence in the WHO Classification of Tumours: a protocol for an evidence gap map of the classification of tumours of the lung. BMJ Open 2022; 12 (10): e061240.

11 Detterbeck FC, Boffa DJ, Kim AW et al. The Eighth Edition Lung Cancer Stage Classification. Chest 2017; 151 (1): 193–203.

12 Welsh JA, Goberdhan DC, O’Driscoll L et al. MISEV2023: An updated guide to EV research and applications. J Extracell Vesicles 2024; 13 (2): e12416.

13 Zhao Z, Wijerathne H, Godwin AK et al. Isolation and analysis methods of extracellular vesicles (EVs). Extracell Vesicles Circ Nucl Acids 2021; 2 (1): 80–103.

14 Welsh JA, Goberdhan DCI, O’Driscoll L et al. Minimal information for studies of extracellular vesicles (MISEV2023): From basic to advanced approaches. J Extracell Vesicles 2024; 13 (2): e12404.

15 Geng X, Tsou JH, Stass SA et al. Utilizing MiSeq Sequencing to Detect Circulating microRNAs in Plasma for Improved Lung Cancer Diagnosis. Int J Mol Sci 2023; 24 (12).

16 Li J, Dhilipkannah P, Holden VK et al. Dysregulation of lncRNA MALAT1 Contributes to Lung Cancer in African Americans by Modulating the Tumor Immune Microenvironment. Cancers (Basel) 2024; 16 (10).

17 Li N, Zhou H, Holden VK et al. Streptococcus pneumoniae promotes lung cancer development and progression. iScience 2023; 26 (2): 105923.

18 Ji P, Diederichs S, Wang W et al. MALAT-1, a novel noncoding RNA, and thymosin beta4 predict metastasis and survival in early-stage non-small cell lung cancer. Oncogene 2003; 22 (39): 8031–8041.

19 Wang J, Tian X, Han R et al. Downregulation of miR-486-5p contributes to tumor progression and metastasis by targeting protumorigenic ARHGAP5 in lung cancer. Oncogene 2014; 33 (9): 1181–1189.

20 Duan J, Cui L, Zhao X et al. Use of Immunotherapy With Programmed Cell Death 1 vs Programmed Cell Death Ligand 1 Inhibitors in Patients With Cancer: A Systematic Review and Meta-analysis. JAMA Oncol 2020; 6 (3): 375–384.

21 Parvez A, Choudhary F, Mudgal P et al. PD-1 and PD-L1: architects of immune symphony and immunotherapy breakthroughs in cancer treatment. Front Immunol 2023; 14: 1296341.

22 Tong C, Wu Y, Wu R. New dimensions of PD-1/PD-L1 inhibitor combination therapy in cancer treatment: current advances and future perspectives. Front Immunol 2025; 16: 1616872.

23 Lin Q, Wang X, Hu Y. The opportunities and challenges in immunotherapy: Insights from the regulation of PD-L1 in cancer cells. Cancer Lett 2023; 569: 216318.

24 Zou W. Immune regulation in the tumor microenvironment and its relevance in cancer therapy. Cell Mol Immunol 2022; 19 (1): 1–2.

25 Racacho KJ, Shiau YP, Villa R et al. The tumor immune microenvironment: implications for cancer immunotherapy, treatment strategies, and monitoring approaches. Front Immunol 2025; 16: 1621812.

26 Xu LF, Wu ZP, Chen Y et al. MicroRNA-21 (miR-21) regulates cellular proliferation, invasion, migration, and apoptosis by targeting PTEN, RECK and Bcl-2 in lung squamous carcinoma, Gejiu City, China. PLoS One 2014; 9 (8): e103698.

27 Huang J, Weng Q, Shi Y et al. MicroRNA-155-5p suppresses PD-L1 expression in lung adenocarcinoma. FEBS Open Bio 2020; 10 (6): 1065–1071.

28 Yang M, Xiao R, Wang X et al. MiR-93-5p regulates tumorigenesis and tumor immunity by targeting PD-L1/CCND1 in breast cancer. Ann Transl Med 2022; 10 (4): 203.

29 Dawidowicz M, Kula A, Mielcarska S et al. miREIA - an immunoassay method in assessment of microRNA levels in tumor tissue-pilot study. The impact of miR-93-5p, miR-142-5p and IFNgamma on PD-L1 level in colorectal cancer. Acta Biochim Pol 2021; 68 (2): 247–254.

